# Altered structural-functional coupling in Parkinson’s disease

**DOI:** 10.1101/2023.01.18.23284750

**Authors:** Zhichun Chen, Guanglu Li, Liche Zhou, Lina Zhang, Jun Liu

**Affiliations:** Department of Neurology and Institute of Neurology, Ruijin Hospital affiliated to Shanghai Jiao Tong University School of Medicine, Shanghai, 200025, China; Department of Neurology, The Second Xiangya Hospital, Central South University, 139 Renminzhong Road, Changsha, 410011, China; Department of Biostatistics, Shanghai Jiao Tong University School of Medicine, Shanghai, 200025, China

**Keywords:** Parkinson’s disease, structural-functional coupling, brain networks, clinical manifestations, genetic variation

## Abstract

**Background:** Structural-functional coupling is abnormally altered in a variety of neuropsychiatric disorders and correlates with clinical symptoms of the patients. The relationships between structural-functional coupling and clinical manifestations of Parkinson’s disease (PD) remain unclear.

**Objective:** The purpose of this study is to investigate whether the structural-functional coupling changes in PD and to explore the clinical implications of this network metric.

**Methods:** Structural 3D T1-weighted imaging, diffusion tensor imaging, and resting-state functional magnetic resonance imaging were performed in 22 control subjects and 34 PD patients. Two types of structural-functional coupling (node coupling and network coupling) were derived from structural and functional images. The associations between structural-functional network coupling and clinical characteristics and genetic variations of 73 PD patients from Parkinson’s Progression Markers Initiative database were analyzed.

**Results:** PD patients exhibited reduced structural-functional node couplings in multiple brain networks compared to controls. Structural-functional node coupling could be shaped by age, sex, and disease severity. In addition, this metric was modified by *TMEM175* rs34311866 and *GPNMB* rs199347, two genetic variants conferring PD risk. In contrast, network coupling was less affected in PD. Particularly, structural-functional network couplings were potential predictors of motor symptoms, non-motor symptoms and pathological markers of cerebrospinal fluid in PD patients. Additionally, structural-functional network coupling was significantly correlated with metrics of network topology.

**Conclusions:** Our findings suggest that structural-functional decoupling is an essential network feature in PD and structural-functional network coupling may serve as a valuable trait-like biomarker for disease burden in PD.

## INTRODUCTION

Understanding the associations between structural connectivity (SC) and functional connectivity (FC) is a basic goal of neuroscience, because it can help us to decode how the white matter networks support human cognition and diverse behaviors (1). With current non-invasive imaging technologies, such as structural magnetic resonance imaging (MRI) and functional MRI, as well as graph theory methods, researchers have found that tight SC-FC couplings exist both in interregional connectivity strength and in network topologic organizations (1). In addition, SC-FC coupling shows age-dependent dynamic changes in normal development and correlated with cognitive functions in youth (2). During aging, SC-FC coupling exhibits a specific pattern characterizing aging-related brain changes (3). Sex is also an important demographic factor shaping SC-FC coupling and it has been shown that females had a stronger SC-FC coupling than males (4). Consistently, Gu *et al* (2021) showed that SC-FC coupling was modified by both age and sex, and was highly heritable within certain networks (5). Besides, gut microbes could also shape the strength of SC-FC coupling in cognition-related regions, such as inferior occipital gyrus, fusiform gyrus, and medial superior frontal gyrus (6).

In the past decade, aberrant SC-FC coupling has been observed in multiple neuropsychiatric diseases, including spastic cerebral palsy (7), schizophrenia (8-11), bipolar disorder (12), attention deficit hyperactivity disorder (13), major depressive disorder (14), epilepsy (15-17), migraine (18), cannabis addiction (19), clinically isolated syndrome (20), multiple sclerosis (21), stroke (22), traumatic brain injury (23), mild cognitive impairment (MCI) (24), Alzheimer’s disease (AD) (24), and Parkinson’s disease (PD) (25). Particularly, even in youth offspring of schizophrenia and bipolar disorder patients, altered SC-FC coupling of long-distance connections has been demonstrated (26). Importantly, SC-FC coupling was found to be associated with clinical manifestations of diseases (8, 13, 24). In addition, SC-FC coupling was correlated with pathological biomarkers in neurodegenerative disease (27). For example, in MCI patients with positive β-amyloid (Aβ) deposition, the level of Aβ pathology and SC-FC coupling was negatively correlated (27), while the correlation between Aβ deposition and SC-FC coupling was positive during AD dementia stage (27).

Previous studies calculated SC-FC coupling defined as the correlation coefficient between non-zero edges of SC and non-zero edges of FC in the whole network (2, 5, 20, 26). The limitation of this type of SC-FC coupling is that it only computes the SC-FC coupling at the network level, but doesn’t calculate the SC-FC coupling at each node. Therefore, SC-FC coupling in the whole network (network coupling) didn’t provide any information on the coupling strength between SC and FC in each region of the brain. Currently, it still lacks a method to compute the SC-FC coupling between non-zero edges of SC and non-zero edges of FC in each node of brain networks, which is referred to as “SC-FC node coupling”.

In PD, Ji *et al* (2019) initially reported decreased SC-FC coupling in the left corticospinal tract of PD patients compared to controls (28). Zarkali *et al* (2021) revealed two organizational gradients to SC-FC decoupling in PD: anterior-to-posterior gradient and unimodal-to-transmodal gradient (25). They also demonstrated that dopaminergic and serotonergic transmission were causally associated with SC-FC decoupling in PD (25). However, these studies didn’t investigate whether SC-FC coupling were altered in different networks of the brain and whether age, sex, and disease severity modified the SC-FC coupling in PD. Additionally, whether SC-FC coupling was associated with motor and non-motor symptoms, and CSF pathological markers in PD was still unexplored. Previously, *TMEM175* rs34311866 and *GPNMB* rs199347 have been reported to be associated with the incidence of PD (29). Additionally, both TMEM175 and GPNMB were found to play a key role in the pathogenesis of PD (30-36). Whether SC-FC coupling was associated with genetic variants conferring risk of PD, such as *TMEM175* rs34311866 and *GPNMB* rs199347, remained unknown. In this study, we examined whether SC-FC coupling was significantly different in different brain networks between PD patients and controls using two SC-FC coupling (network coupling and node coupling) computation methodologies. Then we assessed the effects of age, sex, disease severity, and genetic variants on SC-FC coupling of PD patients. Furthermore, we also explored the associations between SC-FC network coupling and clinical phenotypes of PD patients.

## METHODS

### Study population

The fMRI and demographical data in two cohorts of PD patients and age- and sex-matched controls were obtained. The PD patients (n = 34) and control subjects (n = 22) in cohort 1 were collected from June, 2017 to Sept, 2018 at the Department of Neurology in Ruijin Hospital affiliated to Shanghai Jiao Tong University School of Medicine. The study in cohort 1 was undertaken with the understanding and written consent of each subject and with the approval of the ethics committee of Ruijin Hospital, Shanghai Jiao Tong University School of Medicine, Shanghai, China. PD patients were recruited according to the criteria below: diagnosed as PD according to the current clinical diagnostic criteria of Movement Disorders Society (MDS) (37), aged between 50∼80 years old, excluding other neuropsychiatric disorders, no head injury or surgery history, no MRI signal abnormalities (such as white matter hyperintensities and space-occupying lesions) and MRI artefacts. The motor symptoms were evaluated with MDS Unified Parkinson’s Disease Rating Scale (MDS-UPDRS) (38). Scales used for assessment of non-motor symptoms in cohort 1 include Non-Motor Symptom Questionnaire (NMSQ) (39), Rapid Eye Movement Sleep Behavior Disorder Screening Questionnaire (RBDSQ) (40), Scale for Outcomes in Parkinson disease-Autonomic (SCOPA-AUT) (41), Sniffin Sticks 16-item test (SS-16) (42), Hamilton Depression Rating Scale-17 (HAMD-17) (43), Mini-Mental State Examination (MMSE) (44). The age- and sex-matched control subjects (Age range: 55∼73 years) in cohort 1 were included based on the following criteria: no evidence of significant neurological or psychiatric illnesses during either the structured interview or the formal neurological examination, no first-degree family member with PD, no visually obvious structural abnormalities on their MRI images. The demographic characteristics of PD patients and controls in cohort 1 were shown in Table 1.

**Table 1.** Demographic characteristics for both control and PD patients in Cohort 1

| Characteristic | Control (n = 22) | All patients (n = 34) | Statistic *p* value |
| --- | --- | --- | --- |
| Age (years) | 64.95 ± 4.58 | 64.32 ± 5.03 | *p* > 0.05 |
| Sex (Female/Male) | 13/9 | 15/19 | *p* > 0.05 |
| Education (years) | 10.86 ± 3.73 | 11.71 ± 2.68 | *p* > 0.05 |
| UPDRS-III | 0.55 ± 1.87 | 21.00 ± 9.85 | *p* < 0.0001**** |
| NMSQ | 3.32 ± 2.85 | 7.71 ± 3.56 | *p* < 0.0001**** |
| SS-16 | 12.05 ± 2.01 | 7.62 ± 2.86 | *p* < 0.0001**** |
| RBDSQ | 0.73 ± 1.03 | 5.29 ± 3.71 | *p* < 0.0001**** |
| HAMD-17 | 2.95 ± 4.23 | 3.91 ± 4.20 | *p* > 0.05 |
| SCOPA-AUT | 4.23 ± 4.86 | 10.18 ± 7.57 | *p* < 0.01** |
| MMSE | 27.77 ± 2.62 | 27.79 ± 2.01 | *p* > 0.05 |

Sex ratio was evaluated with Chi-square test, other characteristics were compared using unpaired t-test. UPDRS-III, Unified Parkinson's Disease Rating Scale part III; NMSQ, Non-Motor Symptoms Questionnaire; SS-16, Sniffin Sticks Test; RBDSQ, Rapid-eye-movement Sleep Behavior Disorder Screening Questionnaire; HAMD-17, Hamilton Depression Scale; SCOPA-AUT, Scales for Outcomes in Parkinson's Disease-Autonomic; MMSE, Mini-Mental State Examination.

The PD patients and control subjects in cohort 2 were recruited between Oct, 2012 and Jan, 2017 and were all from Parkinson’s Progression Markers Initiative (PPMI) database. PPMI aims to provide longitudinal imaging and cerebrospinal fluid biomarker datasets for the investigations of PD progression. The methods and protocols of the PPMI have been described previously (45, 46). The inclusion and exclusion criteria of PD patients for cohort 2 also have been published previously (46). The study involved in cohort 2 was approved by the Institutional Review Board of each investigating site. The written informed consents are signed by the study participants and collected by each site investigators. Totally 73 PD subjects in cohort 2 were included for the final analysis in this study. The clinical data of these subjects were all downloaded from PPMI database, including age, sex, disease duration, years of education, Hoehn & Yahr stage, Montreal Cognitive Assessment (MoCA) score (47), SCOPA-AUT score, RBDSQ score, UPDRS-III score, Symbol Digit Modalities Test (SDMT) score, Benton Judgement of Line Orientation (BJLOT) score, and derived scores of Hopkins Verbal Learning Test – Revised (HVLT-R). All the PD subjects in cohort 2 also performed genotyping of *TMEM175* rs34311866 and *GPNMB* rs199347. In addition, almost all the patients in cohort 2 received the examination of iodine-123-labelled ioflupane DAT single photon emission computer tomography. The levels of α-synuclein (α-syn), p-tau, and tau were also available for most of the participants and downloaded from the PPMI database.

Both the participants in cohort 1 and cohort 2 received 3D T1 magnetization-prepared rapid acquisition gradient echo (MPRAGE) imaging, resting-state imaging, and diffusion tensor imaging (DTI). However, because MRI data were not available for most of control participants in cohort 2, they were not included in the neuroimaging analysis.

### Image acquisition

Imaging data acquisition in cohort 1 was performed at the Functional Imaging Centre of the Institutes of Neuroscience of the Chinese Academy of Science in Shanghai using a 3T Siemens Trio MRI scanner (Siemens Healthcare, Malvern, PA) equipped with a 12-channel coil. T1-weighted images were obtained using a 3D MPRAGE sequence: 192 axial slices, Flip Angle = 8°, Voxel size = 1 × 1 × 1 mm^3^, Echo time (TE)/Repetition time (TR)/Inversion time = 4.7 ms/2040 ms/900 ms. Resting-state fMRI data were acquired using gradient echo planar imaging (EPI) sequence: TR = 2000 ms, TE = 28 ms, Flip Angle = 89°, Resolution = 3 × 3 × 3.5 mm. 250 repetitions were acquired over 8 min. Participants were instructed to remain still and awake with their eyes closed during the resting-state fMRI imaging. A 2-dimensional diffusion-weighted, spin-echo, EPI sequence was used for DTI imaging, with diffusion weighting in 28 uniformly distributed directions (b = 1,000 s/mm^2^) and 4 acquisitions without diffusion weighting (b = 0 s/mm^2^): TE /TR = 86.4/13,000 ms, Flip Angle = 90°, Acquisition matrix= 128 × 128, Reconstruction matrix = 256 × 256, Field of View=240 mm, Slice thickness = 3 mm, and Reconstructed voxel size = 1.07 × 1.07 × 3 mm^3^.

The whole-brain structural and functional images in cohort 2 were obtained on 3T Siemens Trio Tim MR system (Siemens Healthcare, Malvern, PA). The detailed MRI technical operations manual can be found at http://www.ppmi-info.org/. 3D T1 structural images were acquired using a MPRAGE sequence with TR = 2300 ms, TE = 2.98 ms, Voxel size = 1 mm^3^, Field of View = 256 mm, Flip Angle =9°, Slice thickness =1.2 mm. The echo planar 2D resting-state images were acquired with TR = 2400 ms, TE = 25 ms, Voxel size = 3.3 mm^3^, Field of View = 222 mm, Flip Angle = 80°, Slice thickness = 3.3 mm. The participants were instructed to rest quietly, keep eyes open, and not fall asleep. The 2D single-shot echo-planar DTI images were acquired with the following parameters: TR= 8,400-8,800 ms, TE = 88 ms, Voxel size = 2 mm^3^, Flip Angle = 90°, Slice thickness = 2 mm, 64 sensitization directions, and a b-value of 1000 s/mm^2^.

### Imaging preprocessing

All the individual images were visually examined for signal dropout or other artifacts before preprocessing. Additionally, DICOM format images were converted into NIFTI format images before the preprocessing of structural and functional images.

Voxel-based morphometry (VBM) was performed to preprocess the 3D T1 images using SPM12 software package (https://www.fil.ion.ucl.ac.uk/spm/software/spm12/) and VBM8 toolbox (http://dbm.neuro.uni-jena.de/vbm8/) (48). Briefly, the 3D T1 images were bias field corrected and normalized to the Montreal Neurological Institute (MNI) template using the diffeomorphic anatomical registration through exponentiated lie algebra (DARTEL) algorithm. Then the normalized images were further segmented into gray matter, white matter, and cerebrospinal fluid (CSF). The processed images were further smoothed before the construction of gray matter covariance network.

In order to preprocess the resting-state images, the first 10 volumes of individual functional image were removed. Slice timing correction was applied to compensate the temporal offsets between slices. In order to keep each part of the brain in the same position, the individual images in all volumes are realigned. In order to adjust the variances of individual brain size, shape and anatomical structure, EPI template was used to convert the image into standard MNI space. An isotropic Gaussian filter(σ = 4mm) was used for spatial smoothing to improve the signal-to-noise ratio and reduce the anatomical variance. The obtained images were further band-pass filtered in low-frequency components (0.01∼0.1HZ). In addition, confounding variables including head motion profiles, CSF signal and white matter signal were regressed to reduce non-neuronal fluctuations. Participants whose head motion frame displacement (FD) > 0.5 mm or head rotation > 2° were excluded. Finally, there was no significant difference in the mean FD values between the study groups.

FMRB Software Library (FSL, https://fsl.fmrib.ox.ac.uk/fsl/fslwiki) was used for the preprocessing of DTI images. The brain extraction tool was used to remove the non-brain tissue and estimate the brain mask. Eddy_correct in FSL was used to correct the eddy current distortions and head motions. Dtifit was used to linearly fit single tensor in single voxel in the brain. Then, the diffusion tensor models were established and independent maps of fractional anisotropy (FA), mean diffusivity, axial diffusivity, radial diffusivity were obtained. Finally, the fnirt and the applywarp commands in FSL were used to normalize the images into the MNI space.

### Network construction

For the construction of functional networks, the Automated Anatomical Labeling (AAL) atlas was utilized to define 90 cortical and subcortical nodes. Then the average time series were extracted for each node and pairwise FC was then measured by computing linear Pearson’s correlation coefficients. This generated a 90 × 90 correlation matrix for each participant. In general, the r-to-z transformation was applied for every correlation matrix to improve the normality of data.

PANDA (https://www.nitrc.org/projects/panda/) was used to construct the white matter network. Ninety cortical and subcortical regions of interest in the AAL atlas were defined as 90 nodes in the white matter network. The whole-brain white matter reconstruction was achieved using deterministic tractography via the Fiber Assignment by Continuous Tracking (FACT) algorithm. For a voxel with FA < 0.2 or a curvature angle > 45°, the fiber tracking process was automatedly terminated. The edge wights (EW), defined as the product of fiber number and FA, was used to define the SC.

In this study, gray matter volume was used as a morphological measure to construct the gray matter covariance network. Detailed methods for the construction of gray matter covariance network have been published previously (49, 50). In brief, AAL atlas was used to define the 90 cortical and subcortical nodes in the gray matter covariance network. The Pearson correlation coefficient between gray matter volumes of each pair of nodes was calculated as the network edge. Correlation coefficients were r-to-z transformed to improve the normality of the data. This produced a 90 × 90 gray matter covariance matrix for each group of participants.

### Node coupling

As mentioned above, the gray matter covariance network (similarity index as node edge), white matter network, and functional network were constructed from structural and functional images. Then the non-zero edge strengths in every type of network were extracted for each pair of nodes. Then, the non-zero edge strengths in the same pair of nodes of 3 types of networks were correlated with each other to compute the node couplings using Pearson’ correlation. As a result, three types of 90 × 90 correlation coefficient matrices were created, including gray matter-functional (GM-Fun) correlation matrix, white matter-functional (WM-Fun) correlation matrix, and gray matter-white matter (GM-WM) correlation matrix. Thus, here we created a new type of coupling, we called it “node coupling”, which calculated the correlation coefficient between edge strength of a node in one type of network and edge strength of the same node in another type of network. Here, two types of SC-FC node coupling were shown. The WM-Fun node coupling calculated the SC-FC node coupling between white matter network and functional network, while the GM-Fun node coupling computed the SC-FC node coupling between gray matter covariance network and functional network. The same method also enabled us to calculate the SC-SC coupling between gray matter covariance network and white matter network.

We also computed the SC-FC node coupling in 5 subnetworks, including default mode network (DMN), basal ganglia network (BGN), executive control network (ECN), sensorimotor network (SMN), and visual (VIS) network. The absolute value of correlation coefficient in each pair of nodes was utilized to measure the strength of node coupling. The detailed computation processes for node coupling were shown in Figure 1.

**Fig. 1.**
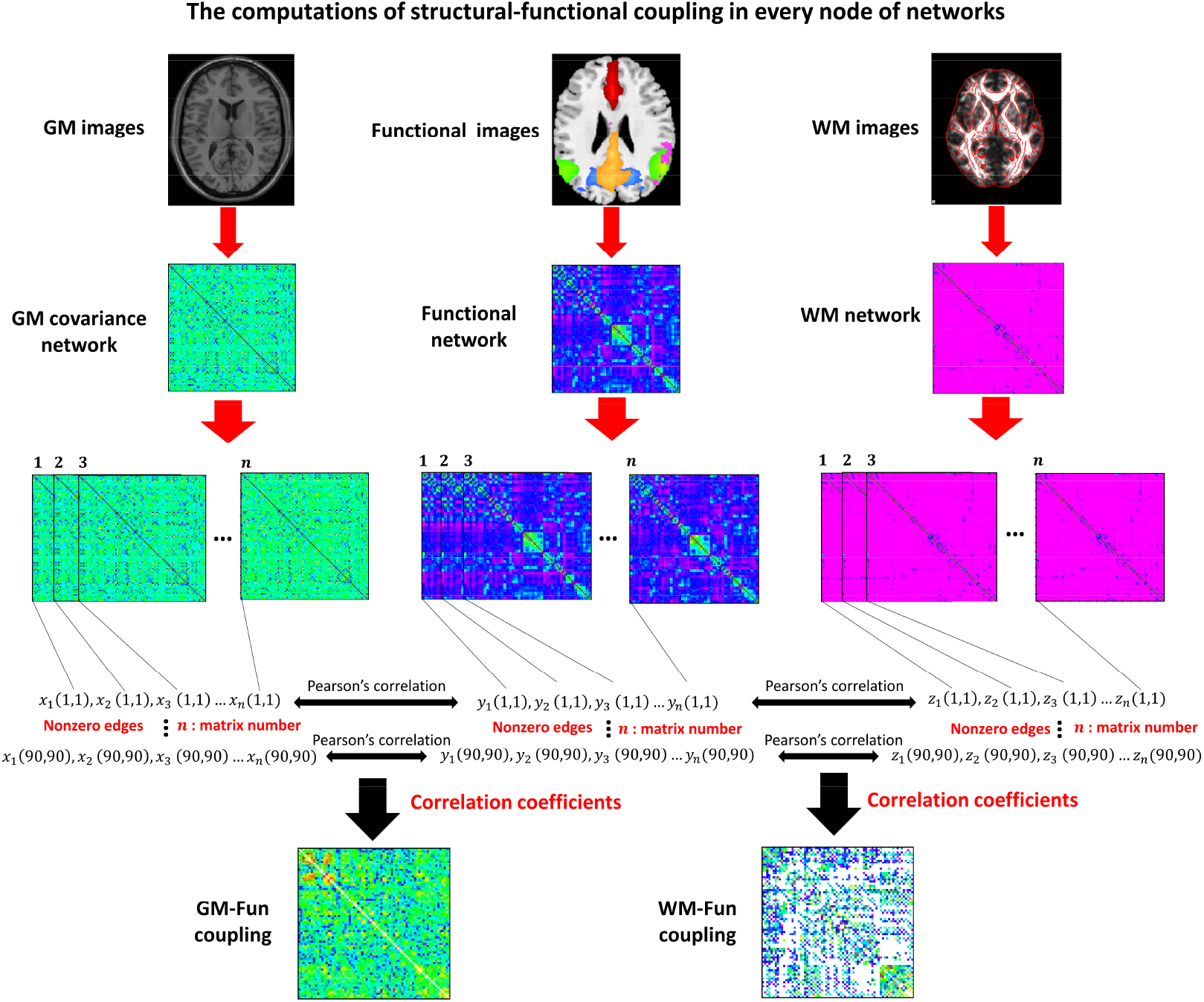
The computation methodology of SC-FC node coupling. The 90 × 90 gray matter covariance network, white matter network, and functional network were extracted from structural and functional images, respectively. Then the non-zero edge strengths in every type of network were extracted for each pair of nodes and correlated with each other to compute their couplings in the same pair of nodes using Pearson’s correlation. This will create three types of 90 × 90 correlation coefficient matrix: gray matter-functional (GM-Fun) correlation matrix, white matter-functional (WM-Fun) correlation matrix, and gray matter-white matter (GM-WM) correlation matrix. The absolute value of correlation coefficient in each pair of nodes was used to measure the strength of node coupling.

### Network coupling

The nonzero edge strengths of functional and structural networks are extracted and correlated to calculate their couplings using Pearson’s correlation. This creates the GM-Fun correlation coefficient and WM-Fun correlation coefficient for each individual, respectively. The GM-Fun correlation coefficient represents the network coupling between the gray matter covariance network and the functional network. The WM-Fun correlation coefficient measures the network coupling between the white matter network and the functional network. Similarly, we also calculate the GM-WM network coupling, which is defined as the correlation coefficient between the gray matter covariance network and the white matter network. The absolute value of correlation coefficient for each individual is used to measure the strength of the network coupling. The computation method for SC-FC network coupling is shown in supplementary Figure 1.

### Graphical network analysis

The GRETNA toolbox (https://www.nitrc.org/projects/gretna/) was utilized to compute topological metrics in gray matter covariance network, functional network, and white matter network. The graphical metrics include assortativity, hierarchy, global efficiency, local efficiency, and small-worldness properties: clustering coefficient (Cp), characteristic path length (Lp), normalized clustering coefficient (γ), normalized characteristic path length (λ), and small worldness (σ). The area under the curve (AUC) for each network metric was derived and used for the correlation analysis between network coupling and graphical metrics. Detailed definitions for each topological metric can be found in previous studies (51, 52).

### Statistical analysis

#### Comparison of clinical characteristics

Differences of clinical characteristics between 2 groups were analyzed using unpaired t-test for continuous variable and Chi-square test for categorical variable. *p* < 0.05 was considered statistically significant.

#### Comparison of coupling strengths

Anderson-Darling test and Shapiro-Wilk test were used for the normality test and revealed that coupling strengths data didn’t pass the normality test (*p* < 0.0001). Differences in means of coupling strengths between 2 groups were analyzed using nonparametric Mann-Whitney U test. For comparisons of coupling strengths among more than 2 groups, nonparametric Kruskal-Wallis test following post hoc Dunn’s test was used. The criteria for significance were: ns (not significant) *p* > 0.05, \**p* < 0.05, \*\**p* < 0.01, \*\*\**p* < 0.001, \*\*\*\**p* < 0.0001.

#### Correlation analysis

The correlations between network couplings (or coupling strengths) and clinical variables or network metrics were assessed with Pearson’s correlation. *p* < 0.05 was considered statistically significant.

## RESULTS

### Demographic data of the study participants

The comparisons of demographical data between control and PD participants in cohort 1 were shown in Table 1. Compared to controls (n = 22), PD patients (n = 34) exhibited higher scores of UPDRS-III (*p* < 0.0001), NMSQ (*p* < 0.0001), RBDSQ (*p* < 0.0001), SCOPA-AUT (*p* < 0.01) and lower scores of SS-16 (*p* < 0.0001). The demographical data in PPMI cohort (cohort 2) was shown in Table 2.

**Table 2.** Demographic characteristics, SBRs, and pathological markers in CSF of PD patients from PPMI cohort.

| Characteristics | All patients<br>(n = 73) | Characteristics | All patients<br>(n = 73) |
| --- | --- | --- | --- |
| Age (years) | 63.49 ± 8.22 | SBR of left caudate | 1.95 ± 0.62 |
| Sex (Female/Male) | 26/47 | SBR of right caudate | 1.94 ± 0.56 |
| Education (years) | 15.00 ± 2.96 | SBR of left putamen | 0.82 ± 0.37 |
| Disease duration (years) | 3.18 ± 2.61 | SBR of right putamen | 0.84 ± 0.35 |
| UPDRS-III | 22.55 ± 11.46 | SBR of left striatum | 2.77 ± 0.92 |
| HY | 1.67 ± 0.53 | SBR of right striatum | 2.78 ± 0.85 |
| Tremor | 4.01 ± 3.68 | SBR of bilateral caudate | 1.95 ± 0.53 |
| RBDSQ | 4.38 ± 2.76 | SBR of bilateral putamen | 0.83 ± 0.30 |
| SCOPA-AUT | 10.90 ± 5.60 | SBR of bilateral striatum | 1.39 ± 0.40 |
| MoCA | 26.45 ± 3.14 | Aβ levels | 848.85 ± 356.54 |
| BJLOT | 12.68 ± 2.01 | α-synuclein levels | 1462.18 ± 608.72 |
| SDMT | 38.84 ± 11.26 | tau levels | 166.22 ± 57.58 |
| HVLT-R: Derived Total Recall | 45.51 ± 13.94 | p-tau levels | 14.60 ± 5.26 |
| HVLT-R: Derived Delayed Recall | 45.53 ± 13.84 | tau/Aβ ratio | 0.21 ± 0.09 |
| HVLT-R: Derived Retention | 47.03 ± 13.23 | p-tau/Aβ ratio | 0.02 ± 0.01 |
| HVLT-R: Derived Discrimination Index | 45.73 ± 12.80 | p-tau/tau ratio | 0.08 ± 0.01 |

Values are expressed as mean ± SD. Abbreviations: Aβ, β-amyloid; HY, Hoehn & Yahr stage; UPDRS-III, Unified Parkinson's Disease Rating Scale Part III; RBDSQ, REM Sleep Behavior Disorder Screening Questionnaire; SCOPA-AUT, Scale for Outcomes in Parkinson's Disease-Autonomic; SDMT, Symbol Digit Modalities Test; BJLOT, Benton Judgement of Line Orientation; MoCA, Montreal Cognitive Assessment; HVLT-R, Hopkins Verbal Learning Test-Revised; SBR, striatal binding ratio; CSF, cerebrospinal fluid.

### Reduced SC-FC node coupling in PD

We firstly examined whether SC-FC coupling was significantly different between control and PD subjects based on the data of cohort 1. We derived two types of SC-FC coupling from structural and functional images of cohort 1. The computation methodologies of these two types of SC-FC coupling have been described in Method as mentioned above and depicted in Figure 1 and supplementary Figure 1. Based on the method shown in Figure 1, we computed SC-FC coupling in every node of the network (referred to as node coupling). Then, we examined whether the distribution of SC-FC couplings among 90 nodes were significantly different between control and PD patients. As shown in Figure 2, the SC-FC node coupling between gray matter covariance network and functional network (GM-Fun coupling) in PD patients was significantly reduced compared to control participants (Fig. 2A; *p* < 0.0001, Mann-Whitney U test). In addition, the node SC-FC coupling between white matter network and functional network (WM-fun coupling) in PD patients was also lower than that of control subjects (Fig. 2B; *p* < 0.0001, Mann-Whitney U test). We also compared the node coupling strength between gray matter covariance network and white matter network and revealed significant reductions of GM-WM coupling in PD patients compared to control (Fig. 2C; *p* < 0.0001, Mann-Whitney U test).

**Fig. 2.**
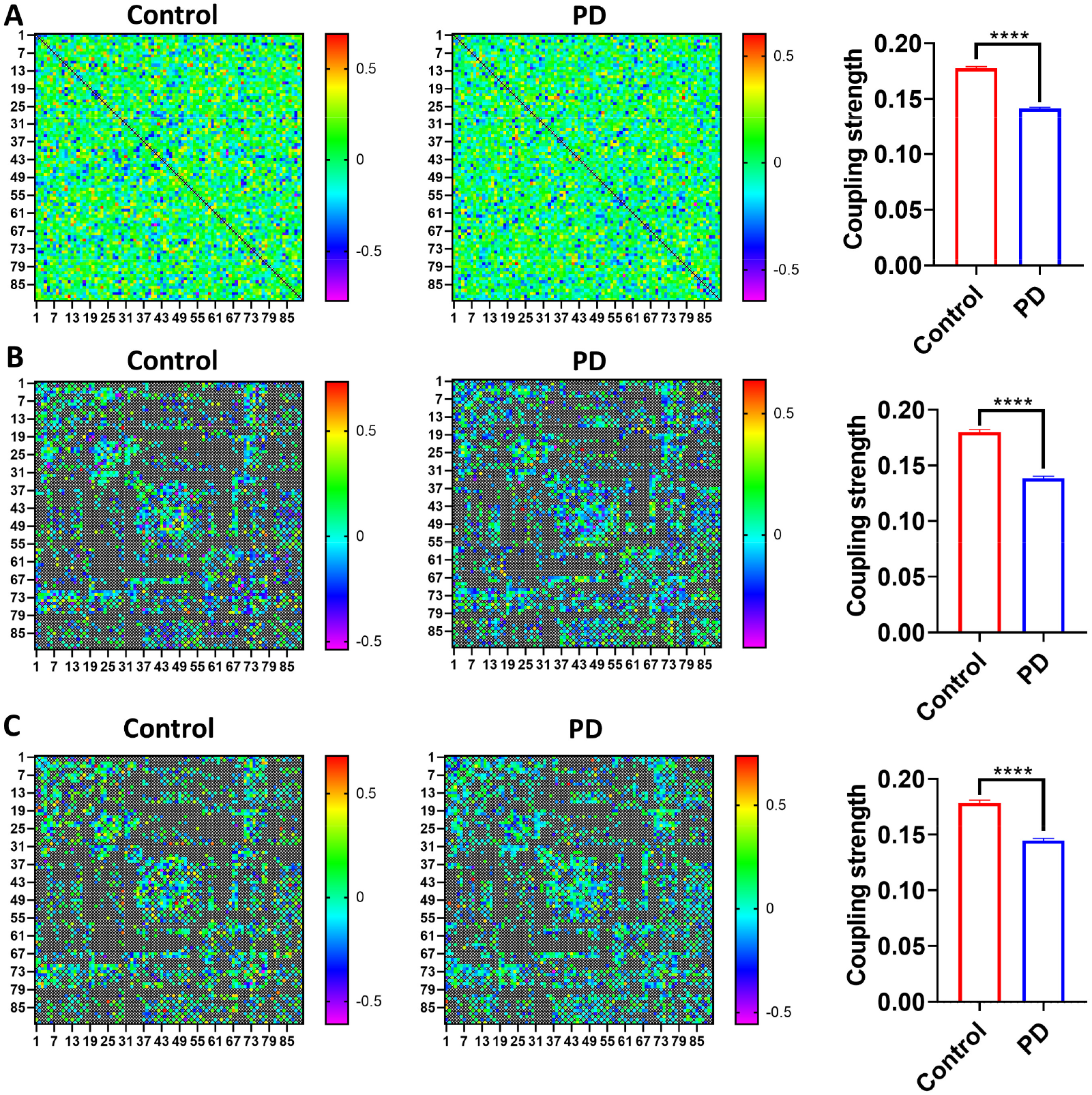
Reduced node coupling strengths in the global network of PD patients. The strengths of GM-Fun node coupling (A), WM-Fun node coupling (B), and GM-WM node coupling (C) were significantly reduced in PD compared to controls. Nonparametric Mann-Whitney U test was used to compare the difference of node coupling strengths between controls and PD patients. \*\*\*\**p* < 0.0001.

We also compared the strengths of node coupling in 5 brain networks. As shown in Figure 3, compared to control subjects, PD patients exhibited reduced GM-Fun node coupling (Fig. 3A-E) in BGN (*p* < 0.01, Mann-Whitney U test), DMN (*p* < 0.05, Mann-Whitney U test), ECN (*p* < 0.01, Mann-Whitney U test), and VIS network (*p* < 0.0001, Mann-Whitney U test) but not in SMN (*p* > 0.05, Mann-Whitney U test) network. Besides, PD patients showed reduced WM-Fun node coupling (Fig. 3F-J) in BGN (*p* < 0.05, Mann-Whitney U test), DMN (*p* < 0.05, Mann-Whitney U test), SMN (*p* < 0.001, Mann-Whitney U test), and VIS (*p* < 0.01, Mann-Whitney U test) network but not in ECN (*p* > 0.05, Mann-Whitney U test) network. Furthermore, we also compared the group difference of GM-WM node coupling (Fig. 3K-O) and found only DMN (*p* < 0.0001, Mann-Whitney U test) network showed reduced GM-WM node coupling in PD patients without changes in other networks.

**Fig. 3.**
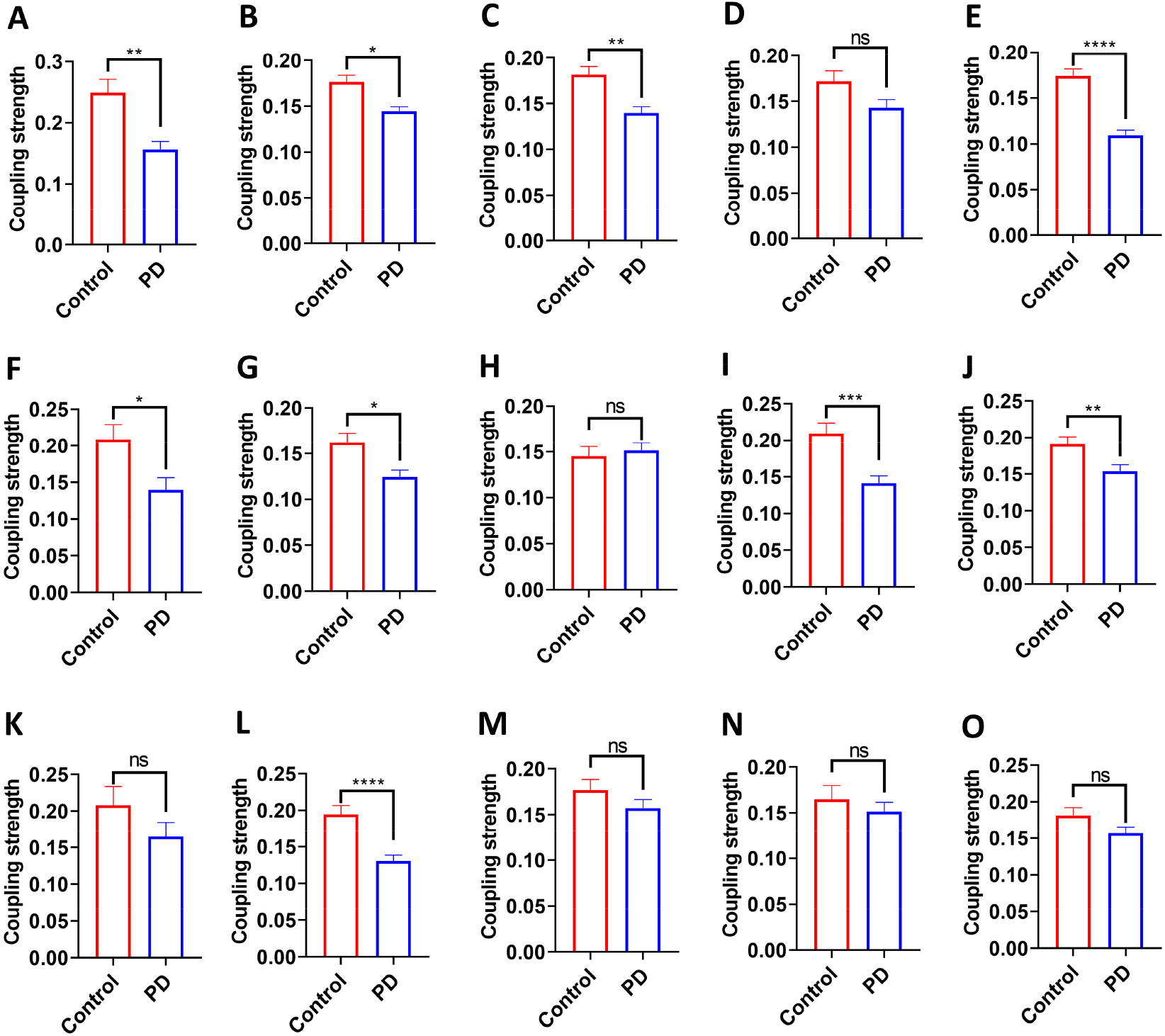
Reduced node coupling strengths in the 5 subnetworks of PD patients. Group differences of GM-Fun node coupling strengths in BGN (A), DMN (B), ECN (C), SMN (D), and VIS (E). Group differences of WM-Fun node coupling strengths in BGN (F), DMN (G), ECN (H), SMN (I), and VIS (J). Group differences of GM-WM node coupling strengths in BGN (K), DMN (L), ECN (M), SMN (N), and VIS (O). Nonparametric Mann-Whitney U test was used to compare the difference of node coupling strengths between controls and PD patients. \**p* < 0.05, \*\**p* < 0.01, \*\*\**p* < 0.001, \*\*\*\**p* < 0.0001.

Then we computed the second type of SC-FC coupling (referred to as network coupling) in both control and PD participants using the methodology presented in supplementary Figure 1 and found that GM-Fun network coupling, WM-Fun network coupling, and GM-WM network coupling were not significantly different between PD patients and control subjects (all *p* > 0.05, data not shown).

Because most control participants in cohort 2 have no MRI data, we can’t validate above significant difference of SF-FC coupling between control and PD participants in cohort 2.

### SC-FC coupling varied among different network types

The SC-FC coupling was derived from the structural and functional images of 73 PD patients in PPMI cohort. For node couplings, there were no significant differences among the strengths of the GM-Fun node coupling, WM-Fun node coupling, and GM-WM node coupling (all *p* > 0.05, data not shown). For network couplings, WM-Fun network coupling was significantly greater than GM-Fun and GM-WM network coupling in the whole network (*p* < 0.0001; Supplementary Fig. 2A). Interestingly, WM-Fun network coupling was also much higher than GM-Fun and GM-WM network coupling in 4 subnetworks, including DMN (*p* < 0.0001), ECN (*p* < 0.0001), SMN (*p* < 0.0001), and VIS (*p* < 0.0001), but not in BGN (Supplementary Fig. 2B-F). We also compared the strengths of network coupling among different networks. For WM-Fun network coupling, WM-Fun network coupling in BGN was much lower than that of other subnetworks (Supplementary Fig. 2G), including DMN (*p* < 0.0001), ECN (*p* < 0.0001), SMN (*p* < 0.0001), and VIS network (*p* < 0.0001). For GM-Fun network coupling, GM-Fun network coupling in DMN was lower than that of other networks (Supplementary Fig. 2H), including BGN (*p* < 0.0001), ECN (*p* < 0.0001), SMN (*p* < 0.0001), and VIS network (*p* < 0.001). In addition, we also revealed that GM-WM network coupling in DMN was much lower than that of other subnetworks (Supplementary Fig. 2I), such as BGN (*p* < 0.05) and SMN (*p* < 0.001). We also explored whether the coupling strengths in 5 subnetworks could predicted the coupling strengths of the whole network. we found WM-Fun network coupling strength in DMN was significantly correlated with WM-Fun network coupling strength in the whole network (r = 0.44, *p* = 0.0001; Supplementary Fig. 2J). GM-WM network coupling strength in VIS was significantly correlated with GM-WM network coupling strength in the whole network (r = 0.40, *p* = 0.0005; Supplementary Fig. 2K).

### Effects of age, sex and disease severity on node couplings

Because we found SC-FC node coupling was significantly altered in PD patients, we further evaluate the effects of age, sex, and disease severity on SC-FC node coupling of PD patients. In 73 PD patients from PPMI cohort, GM-Fun node coupling, WM-Fun node coupling, and GM-WM node coupling in PD patients with age ≥60 (n = 48) was significantly lower than that of patients with age < 60 (n = 25; all *p* < 0.0001; Supplementary Fig. 3A-C). Consistently, GM-Fun node coupling (Supplementary Fig. 3D), WM-Fun node coupling (Supplementary Fig. 3E), and GM-WM node coupling (Supplementary Fig. 3F) in 5 subnetworks also tended to be lower in PD patients with age ≥ 60 than that of patients with age < 60. Compared to male patients (n = 47), female patients (n = 26) exhibited higher GM-Fun node coupling (*p* < 0.0001; Fig. 4A), WM-Fun node coupling (*p* < 0.0001; Fig. 4B), and GM-WM node coupling in the whole network (*p* < 0.0001; Fig. 4C). In addition, female patients were also shown to have higher GM-Fun node coupling (Fig. 4D), WM-Fun node coupling (Fig. 4E), and GM-WM node coupling (Fig. 4F) in 5 subnetworks. Compared to patients with Hoehn & Yahr stage ≤1 (n = 23), patients with Hoehn & Yahr stage ≥2 (n = 50) displayed lower GM-Fun node coupling, WM-Fun node coupling, and GM-WM node coupling in the whole network (all *p* < 0.0001; Supplementary Fig. 4A-C). Consistently, GM-Fun node coupling (Supplementary Fig. 4D), WM-Fun node coupling (Supplementary Fig. 4E), and GM-WM node coupling (Supplementary Fig. 4F) in 5 subnetworks were also prone to be lower in PD patients with Hoehn & Yahr stage ≥2 compared to patients with Hoehn & Yahr stage ≤1.

**Fig. 4.**
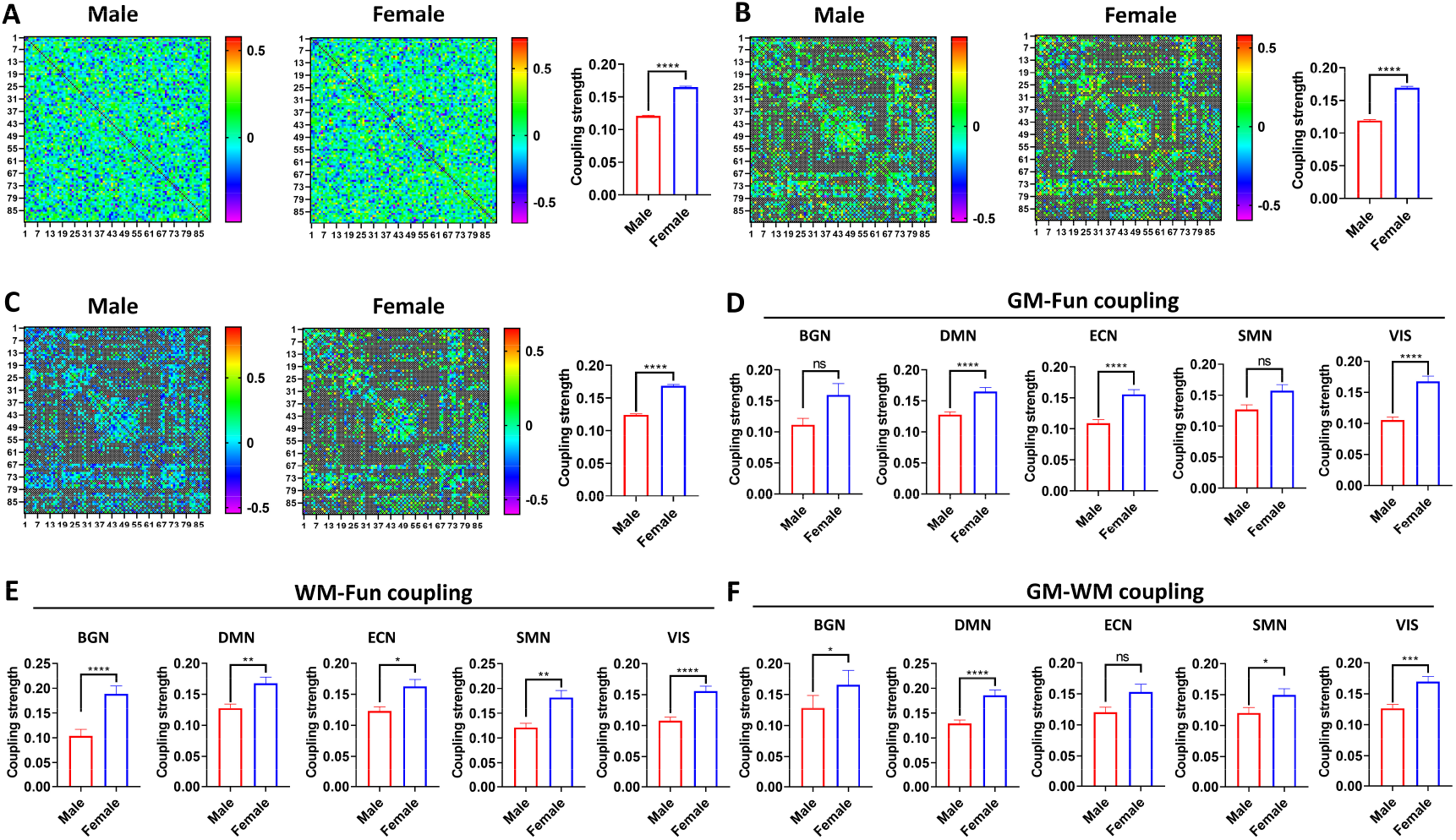
Female patients exhibited higher node coupling strengths compared to male patients. Enhanced strengths of GM-Fun node coupling (A), WM-Fun node coupling (B), and GM-WM node coupling (C) in global network of female patients (n = 26) compared to male patients (n = 47). Group differences in the strengths of GM-Fun node coupling (D), WM-Fun node coupling (E), and GM-WM node coupling (F) in 5 subnetworks. Nonparametric Mann-Whitney U test was used to compare the difference of node coupling strengths between male and female patients. \**p* < 0.05, \*\**p* < 0.01, \*\*\**p* < 0.001, \*\*\*\**p* < 0.0001.

GM-Fun network coupling, WM-Fun network coupling, and GM-WM network coupling was not significantly different between patients with age < 60 and patients with age ≥ 60 (data not shown). Compared to female patients, male patients exhibited lower strength of GM-Fun network coupling in ECN (*p* < 0.01; Supplementary Fig. 5A) and lower strength of WM-Fun network coupling in the whole network (*p* < 0.05; Supplementary Fig. 5B) and SMN (*p* < 0.01; Supplementary Fig. 5C). Compared to patients with Hoehn & Yahr stage ≤1, patients with Hoehn & Yahr stage ≥2 showed higher WM-Fun network coupling in BGN (*p* < 0.01; Supplementary Fig. 5D) and lower GM-WM network coupling in BGN (*p* < 0.05; Supplementary Fig. 5E).

### Genetic modifications of SC-FC coupling

Because brain networks were significantly modified by genetic variations (53-56), we examined whether SC-FC coupling was shaped by genetic variants conferring a risk of PD. We found *TMEM175* rs34311866 T-carriers (n = 25) exhibited higher GM-Fun node coupling (*p* < 0.0001; Fig. 5A), WM-Fun node coupling (*p* < 0.0001; Fig. 5B), and GM-WM node coupling (*p* < 0.0001; Fig. 5C) compared to CC carriers (n = 48). *GPNMB* rs199347 GG carriers (n = 11) showed higher GM-Fun node coupling (both *p* < 0.0001; Fig. 5D), higher WM-Fun node coupling (both *p* < 0.0001; Fig. 5E), and higher GM-WM node coupling (both *p* < 0.0001; Fig. 5F) compared to AA (n = 26) and AG carriers (n = 36). In contrast, we found both *TMEM175* rs34311866 and *GPNMB* rs199347 had no significant effects on GM-Fun network coupling, WM-Fun network coupling, and GM-WM network coupling (all *p* > 0.05; data not shown).

**Fig. 5.**
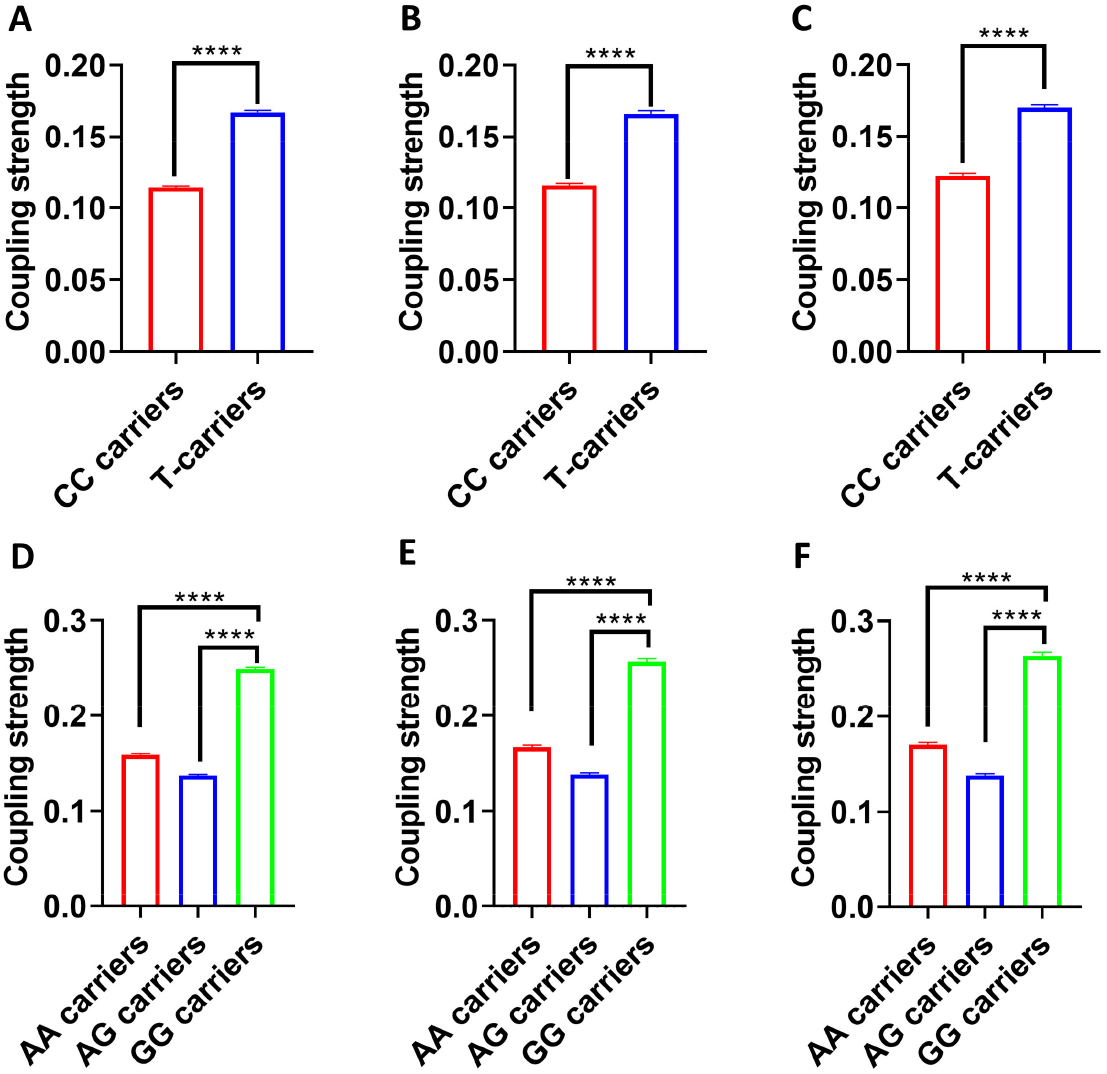
*TMEM175* rs34311866 and *GPNMB* rs199347 modified node coupling strengths in PD patients. *TMEM175* rs34311866 T-carriers (n =25) showed higher strengths of GM-Fun node coupling (A), WM-Fun coupling (B), and GM-WM node coupling (C) in the global network compare to CC carriers (n = 48). *GPNMB* rs199347 GG carriers (n = 11) displayed enhanced strengths of GM-Fun node coupling (D), WM-Fun coupling (E), and GM-WM node coupling (F) in the global network compare to AA carriers (n = 26) and AG carriers (n = 36). Nonparametric Mann-Whitney U test was used to compare the difference of node coupling strengths between *TMEM175* rs34311866 T-carriers and CC carriers. Nonparametric Kruskal-Wallis test following post hoc Dunn’s test was used to compare the difference of node coupling strengths among *GPNMB* rs199347 GG carriers, GA carriers, and AA carriers. \*\*\*\**p* < 0.0001.

### SC-FC coupling and motor symptoms and non-motor symptoms

Next, we examined whether SC-FC network coupling was correlated with clinical variables of PD patients. We found WM-Fun network coupling in BGN was positively correlated with UPDRS-III scores (r = 0.31, *p* < 0.01; Fig. 6A) and SCOPA-AUT scores (r = 0.30, *p* < 0.01; Fig. 6B), but negatively correlated with Derived Delayed Recall scores (r = -0.28, *p* < 0.05; Fig. 6C) and Derived Retention scores (r = -0.30, *p* < 0.05; Fig. 6C) of HVLT-R. WM-Fun network coupling in DMN was positively correlated with Derived Delayed Recall scores (r = 0.27, *p* < 0.05; Fig. 6D) and Derived Retention scores (r = 0.25, *p* < 0.05; Fig. 6D) of HVLT-R. GM-Fun network coupling in ECN was negatively correlated with UPDRS-III scores (r = - 0.32, *p* < 0.01; Fig. 6E) and positively correlated with Derived Delayed Recall scores (r = 0.29, *p* < 0.05; Fig. 6F) and Derived Retention scores (r = 0.28, *p* < 0.05; Fig. 6F) of HVLT-R.

**Fig. 6.**
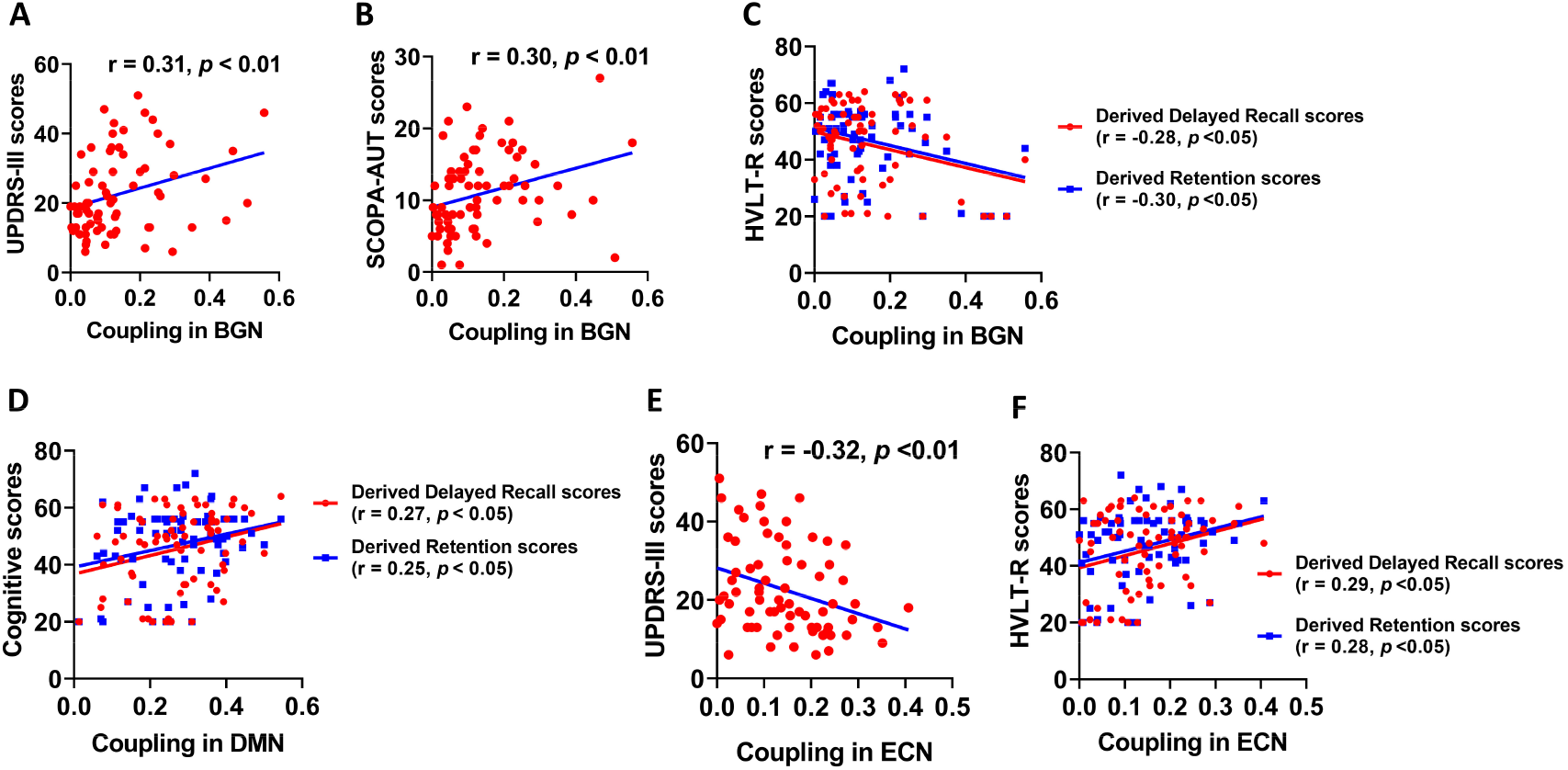
SC-FC network couplings predicted clinical motor and non-motor symptoms. WM-Fun network coupling in BGN was positively corelated with UPDRS-III scores (A) and SCOPA-AUT scores (B) but negatively correlated with Derived Delayed Recall and Derived Retention scores of HVLT-R (C). GM-Fun network coupling in ECN was negatively corelated with UPDRS-III scores (D) but positively correlated with Derived Delayed Recall and Derived Retention scores of HVLT-R (E). WM-Fun network coupling in DMN was positively corelated with Derived Delayed Recall and Derived Retention scores of HVLT-R (F). Pearson’s correlation was used to analyze the relationships between SC-FC network coupling and scores of clinical scales.

### SC-FC network coupling and pathological proteins in CSF

We found WM-Fun network coupling was not significantly correlated with α-synuclein, tau, and p-tau levels in CSF. In contrast, GM-Fun network couplings in the whole network and SMN were negatively correlated with α-synuclein (r =-0.24, *p* < 0.05 and r = -0.35, *p* < 0.01; Fig. 7A-B) and tau levels (r =-0.37, *p* < 0.01 and r = - 0.33, *p* < 0.01; Fig. 7C-D). In addition, GM-Fun network coupling in the whole network and VIS was negatively correlated with p-tau levels (r =-0.37, *p* < 0.01 and r = -0.27, *p* < 0.05; Fig. 7E-F).

**Fig. 7.**
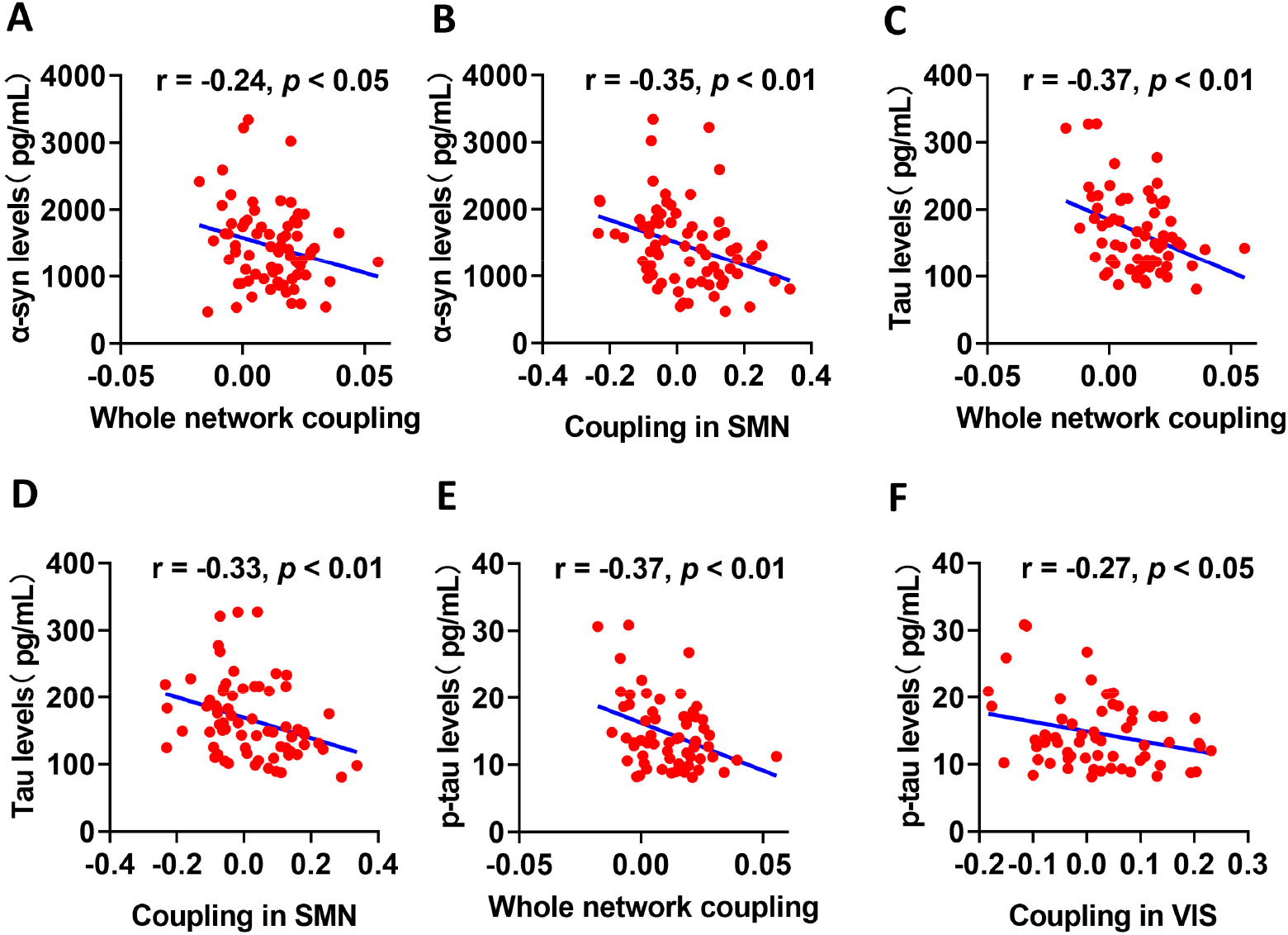
GM-Fun network couplings predicted the burden of CSF pathological proteins. GM-Fun coupling in the whole network and SMN were negatively correlated with α-synuclein (A and B) and tau (C and D) levels in CSF. GM-Fun coupling in the whole network and VIS were negatively correlated with p-tau (E and F) levels in CSF. Pearson’s correlation was used to analyze the relationships between SC-FC network coupling and the levels of CSF pathological proteins.

### SC-FC network coupling and graphical metrics

As a different type of network metric, whether SC-FC couplings are reliable measures of brain networks remains unknown. Thus, we explored whether SC-FC coupling was associated with other network metrics, such as global efficiency and small-worldness properties. As shown in Figure 8, we revealed that WM-Fun network coupling in the whole network was negatively correlated with assortativity (r = -0.24, *p* < 0.05; Fig. 8A) and positively correlated with hierarchy (r = 0.23, *p* < 0.05; Fig. 8B), global efficiency (r = 0.28, *p* < 0.05; Fig. 8C), small-worldness Lp (r = 0.26, *p* < 0.05; Fig. 8D), small-worldness γ (r = 0.25, *p* < 0.05; Fig. 8E), and small-worldness σ (r = 0.26, *p* < 0.05; Fig. 8F) in functional network. Besides, WM-Fun network coupling in the whole network was positively correlated with hierarchy (r = 0.26, *p* < 0.05; Fig. 8G), global efficiency (r = 0.29, *p* < 0.05; Fig. 8H) and negatively correlated with small-worldness Lp (r = -0.30, *p* < 0.01; Fig. 8I), small-worldness γ (r = -0.27, *p* < 0.05; Fig. 8J), small-worldness λ (r = -0.29, *p* < 0.05; Fig. 8K), and small-worldness σ (r = -0.25, *p* < 0.05; Fig. 8L) in white matter network.

**Fig. 8.**
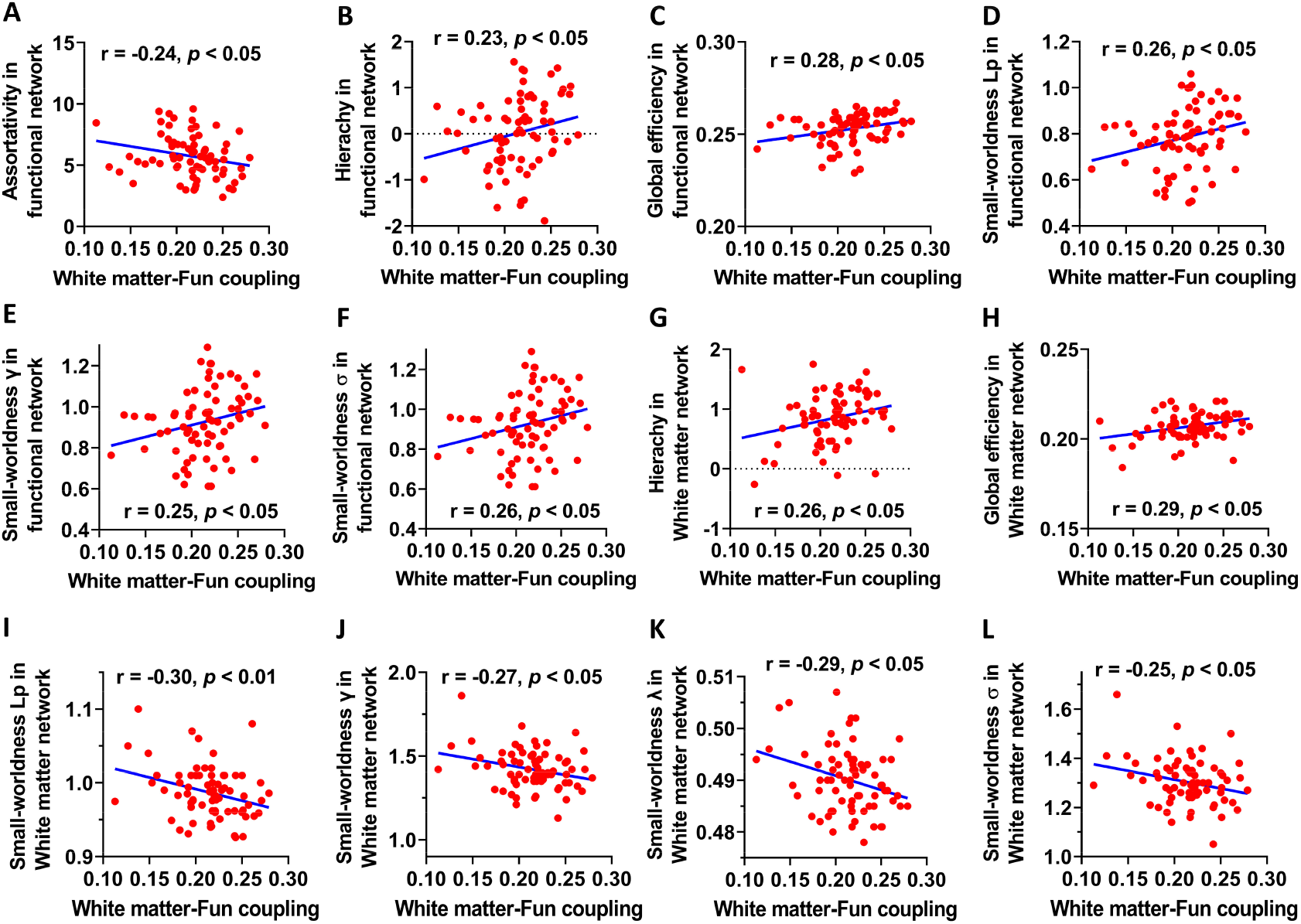
WM-Fun coupling in the whole network was correlated with topological measures in both functional network and white matter network. WM-Fun coupling in the whole network was negatively correlated with assortativity (A) and positively correlated with hierarchy (B), global efficiency (C), small-worldness Lp (D), small-worldness γ (E), and small-worldness σ (F) in functional network. WM-Fun network coupling in the whole network was positively correlated with hierarchy (G), global efficiency (H) and negatively correlated with small-worldness Lp (I), small-worldness γ (J), small-worldness λ (K), and small-worldness σ (L) in white matter network. Pearson’s correlation was used to analyze the relationships between SC-FC network coupling and the graphical metrics of functional network and white matter network.

## DISCUSSION

In this study, we demonstrated that SC-FC node couplings in the whole network were significantly reduced in PD patients, though SC-FC network couplings were not significantly different. Specially, BGN, DMN, and ECN showed both reduced GM-Fun and WM-Fun node couplings in PD patients compared to controls. We found age, sex, and disease severity modified SC-FC node coupling in PD patients, characterized by lower SC-FC node coupling for much older, male, and more severe patients. In addition, we revealed that SC-FC node coupling were shaped by recently identified genetic variants: *TMEM175* rs34311866 and *GPNMB* rs199347. In PPMI cohort, we found SC-FC network coupling in BGN were positively correlated with UPDRS-III scores and negatively correlated with verbal memory performance, while SC-FC network coupling in ECN network were negatively correlated with UPDRS-III scores and positively correlated with verbal memory performance. Additionally, GM-Fun network couplings were negatively correlated with the levels of pathological markers in CSF, such as α-synuclein, tau, and p-tau. Finally, we also revealed that WM-Fun network coupling in the whole network was significantly correlated with topological metrics of functional network and white matter network. Therefore, our study provides a comprehensive analysis on the SC-FC couplings and the associations between SC-FC coupling and clinical characteristics in PD.

There is a consensus in the neuroscience that cognitive functions, behaviors, and FC are supported by brain structural architecture including both white matter structure and gray matter structure (1, 3). SC-FC coupling evaluates the dependence of functional network on structural network and is intimately correlated with cognitive function (1, 3, 21). SC-FC couplings were generally reduced in neuropsychiatric diseases (12, 20, 23, 25); however, increased SC-FC coupling has been found in schizophrenia and epilepsy (8, 17). In current study, we proposed a new type of SC-FC coupling, we called it “node coupling”, which computes the correlation coefficients between SC and FC in each node pair (Fig. 1). This type of SC-FC coupling was not described in previous studies and helped us to evaluate node-level of SC-FC coupling. We found that SC-FC node couplings were significantly reduced in PD patients indicating the reduction of dependence of FC on SC in PD. The reduction of SC-FC node coupling in the whole network of PD patients was companied by the decreases of SC-FC node couplings in some subnetworks, such as BGN, DMN, and VIS, thus our results suggest that SC-FC decoupling was a universal phenomenon in PD patients. Our new computation methodology for SC-FC coupling provides new insights into the disruptions of structure-function associations in PD (25, 28). To differentiate the SC-FC node coupling from previous reported SC-FC coupling, we called the latter “network coupling” which computes the correlation coefficients between SC and FC at the network level. Compared to the node coupling, the network coupling in the whole network was not significantly different between controls and PD patients although the WM-Fun network coupling in ECN was significantly reduced in PD patients. These data indicate that node coupling and network coupling are two different types of coupling metrics, which may provide complimentary information about the relationships between brain structure and function.

Previous studies utilized the SC derived from diffusive MRI to calculate SC-FC coupling; however, it only computes the coupling between white matter connectivity and FC (WM-Fun coupling). In this study, we created a new type of SC-FC coupling, the coupling between gray matter similarity index (49, 50) and FC (GM-Fun coupling). The gray matter covariance network is a new type of structure network assessing the spatial similarity of gray matter structure in the networks, which indirectly reflects the SC between different anatomical regions (50). According to the axon tension theory proposed by van Essen (1997), axons between connected brain regions produce a mechanical force, creating a tension that pulls connected areas together, whereas brain regions that are not directly connected would drift apart (57). This axon tension hypothesis has been demonstrated in monkey and human brains (57-60). Therefore, gray matter covariance network could also be used to calculate SC-FC coupling, which is similar to the SC-FC coupling derived from white matter network and functional network. Compared to WM-Fun coupling, the strength of GM-Fun coupling was much lower in most of brain networks except BGN, thus, white matter connectivity matches much better with FC than gray matter similarity index. In our study, similar to WM-Fun node coupling, we found GM-Fun node coupling was also significantly reduced in PD patients, supporting that GM-Fun coupling showed consistent changes with WM-Fun coupling in PD. Taken together, we propose that GM-Fun coupling is a new type of SC-FC coupling deserved to be investigated in future.

Previous studies demonstrated that age and sex modified SC-FC coupling (2, 4, 5), which was supported by our current study. We found SC-FC node coupling was much lower in patients with age ≥ 60 compared to that of patients with age < 60, indicates that older age may be associated with a decline of SC-FC node coupling in PD patients. It should be noted that age-dependent decline of SC-FC node coupling exist in both GM-Fun coupling and WM-Fun coupling. A previous study reported that female subjects exhibited higher SC-FC node coupling compared to male subjects (4), which was also supported by our study showing that female patients displayed both higher GM-Fun node coupling and WM-Fun node coupling in PD. The results from recent studies and our study suggest that females have stronger relationships between SC and FC. These findings also imply that network integrity in female patients may be less impaired compared to male patients, which was consistent with the results reported by recent studies showing that female patients exhibited milder motor and non-motor symptoms and better cognitive functions compared to male patients (61-64). The mechanisms underlying the enhanced SC-FC coupling in females are still unknown and need to be further investigated. We found patients with Hoehn & Yahr stage ≥2 exhibited much lower SC-FC node coupling compared to patients with Hoehn & Yahr stage ≤ 1, supporting that worse disease severity was associated with much lower SC-FC coupling in PD. The effects of disease severity on SC-FC coupling suggest that SC-FC node coupling was a potential biomarker for disease burden in PD.

A recent study has demonstrated that SC-FC coupling was highly heritable within certain networks (5). Our study also supports this notion by showing that PD-associated risk loci modified SC-FC node coupling in PD patients. Both *TMEM175* rs34311866 and *GPNMB* rs199347 have been shown to confer a risk of PD and have been found to participate in the pathophysiological processes of PD (29-31, 36). We found *TMEM175* rs34311866 T-carriers had higher SC-FC node coupling compared to CC carriers, which was consistent with milder motor and non-motor symptoms in *TMEM175* rs34311866 T-carriers (Supplementary Fig. 6). Wie *et al* (2021) reported that *TMEM175* deficiency leads to dopaminergic neurodegeneration and impairment in motor function in mice and a loss-of-function of *TMEM175* is associated with exacerbation of cognitive and motor decline in PD patients (30). Additionally, *TMEM175* deficiency also resulted in lysosomal over-acidification, disrupted proteolytic activity, and enhanced α-synuclein aggregation in vivo (31). In contrast, a recent study showed that enhancement of TMEM175 function inhibits mitophagy, impairs mitochondrial homeostasis, increases reactive oxygen species, and aggravates symptoms in a 1-methyl-4-phenyl-1,2,3,6-tetrahydropyridine (MPTP) mouse model of PD (32). Thus, both loss-of-function and gain-of-function of *TMEM175* was associated with neurodegeneration in PD. *GPNMB* rs199347 (A > G) was associated a reduced risk of PD (29). Recently, GPNMB has been shown to internalize α-synuclein fibrils and promote α-synuclein pathology, thus, GPNMB is harmful and exacerbates neurodegeneration in PD (36). Consistently, the level of GPNMB was increased in the biofluid of PD patients and positively correlated with UPDRS-III scores (36). Because GG carriers have reduced GPNMB levels compared to AA carriers (36), GG genotype is protective, which is consistent with our findings that GG carriers displayed higher SC-FC node coupling compared to AA carriers in PD. The protective effects of *GPNMB* rs199347 G allele in PD was also supported by our recent study showing that G allele of *GPNMB* rs199347 specifically enhances the activity of sensorimotor network and the information transfer efficiency of functional network (65). To sum, our findings support that SC-FC node coupling was significantly shaped by PD-associated genetic variations.

We found SC-FC network coupling in BGN is positively correlated with UPDRS-III scores whereas SC-FC network coupling in ECN is negatively correlated with UPDRS-III scores. Additionally, SC-FC network coupling in BGN is negatively correlated with verbal memory performance whereas SC-FC network couplings in ECN and DMN are positively correlated with verbal memory performance. These results indicate that SC-FC network coupling in BGN network predicted worse disease severity while SC-FC network coupling in ECN predicted milder disease burden in PD. Future studies will be required to investigate the value of SC-FC network coupling as potential biomarkers for the monitoring of disease progression in PD.

The associations between SC-FC network coupling and pathological markers in neurodegenerative diseases remain unclear. In MCI and AD patients, SC-FC network coupling is correlated with Aβ deposition (27). We found GM-Fun network coupling was negatively correlated with the levels of α-synuclein, tau, and p-tau in CSF. These findings indicate that patients with negative GM-Fun network coupling may have much higher levels of α-synuclein, tau, and p-tau while patients with positive GM-Fun network network coupling have much lower levels of α-synuclein, tau, and p-tau in CSF. Because both tau and p-tau are significantly correlated with neurodegeneration and cognitive deficits (66-68), our results support that GM-Fun network couplings predict neurodegenerative biomarkers in CSF. Future studies are needed to explore the merit of GM-Fun network coupling for the prediction of CSF pathological markers in PD.

As a different type of network metric, we found WM-Fun network coupling in the whole network was significantly correlated with graphical network metrics of functional network and white matter network. These results indicate that WM-Fun network coupling is reliable network metric reflecting the topology of brain functional and structural networks. In both functional network and white matter network, both hierarchy and global efficiency were positively correlated with WM-Fun network coupling, which indicates that WM-Fun network coupling in the whole network reflects efficiency of information transfer in brain networks. Interestingly, the WM-Fun network coupling was positively correlated with small-worldness properties in functional network whereas negatively correlated with small-worldness properties in white matter network. These results support that WM-Fun network coupling may also be a potential measure of small-world topology for brain networks. Taken together, these findings suggested that WM-Fun network coupling was a solid metric for the measurement of brain network topology.

## CONCLUSIONS

Overall, our study demonstrated that SC-FC node couplings were significantly reduced in PD patients compared to controls. In addition, SC-FC node couplings were shaped by age, sex, disease severity, and genetic loci conferring a risk of PD. Furthermore, SC-FC network couplings were potential imaging biomarkers for clinical symptoms, CSF pathology, and brain network topology. These findings strongly support further investigations on the role of SC-FC coupling in the diagnosis of neurological disease and monitoring of disease progression.

## Data Availability

Clinical and imaging data in cohort 2 were obtained from the Parkinson's progression markers initiative (PPMI) database (www.ppmiinfo.org/data). All data produced in the present study are available upon reasonable request to the authors.

http://www.ppmi-info.org/data

## ACKNOWLEDGMENTS

Data in cohort 2 were obtained from the Parkinson’s Progression Markers Initiative (PPMI) database (www.ppmiinfo.org/data). We thank the share of PPMI data by all the PPMI study investigators. For up-to-date information on the study, visit www.ppmiinfo.org. PPMI – a public-private partnership – is funded by the Michael J. Fox Foundation for Parkinson’s Research and funding partners, which can be found at www.ppmiinfo.org/fundingpartners.

## AUTHOR CONTRIBUTIONS

Zhichun Chen, Conceptualization, Formal analysis, Visualization, Methodology, Writing - original draft, Writing - review and editing; Guanglu Li, Data curation, Formal analysis, Visualization; Liche Zhou, Data curation, Formal analysis, Investigation; Lina Zhang, Formal analysis, Investigation, Methodology; Jun Liu, Conceptualization, Supervision, Funding acquisition, Writing - original draft, Project administration, Writing- review and editing.

## SUPPLEMENTAL MATERIAL

The supplementary materials include 6 supplementary figures.

