## Supplemental file for "Altered structural-functional coupling in Parkinson’s disease"

### Supplementary Material

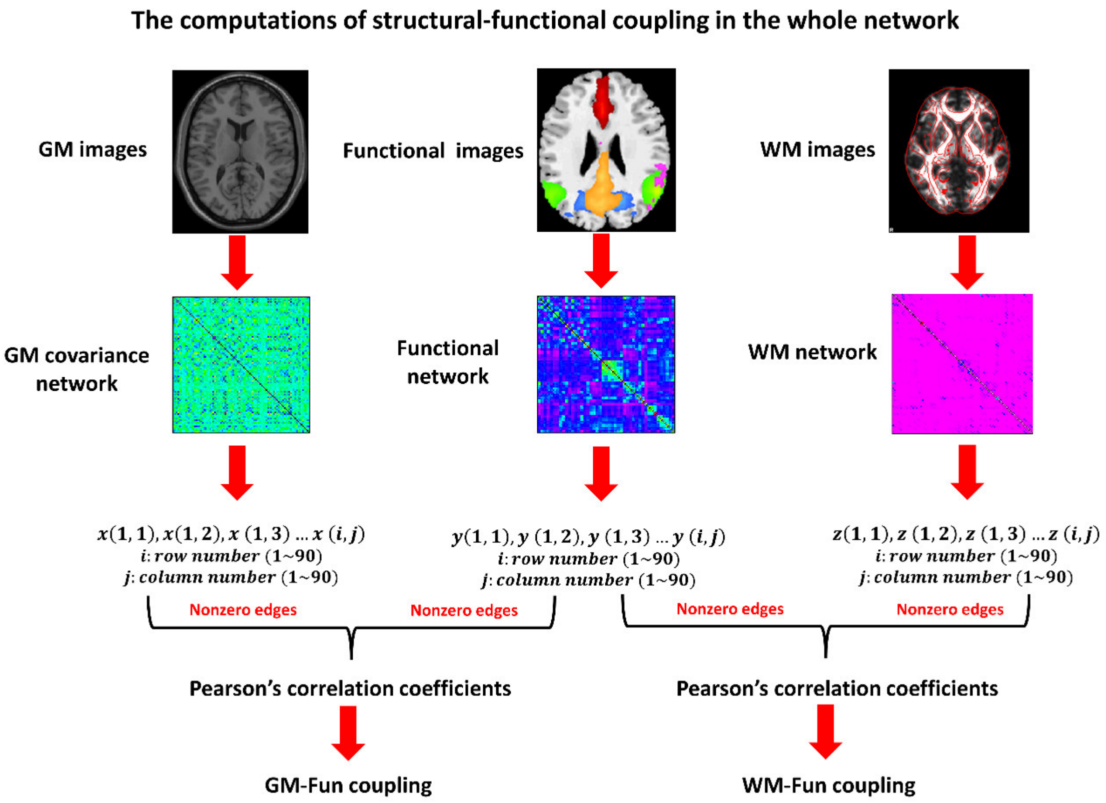


Supplementary Fig. 1. The computation methodology of SC-FC network coupling. The non-zero edge strengths were extracted for both functional and structural network and correlated with each other to compute their couplings. This will create a GM-Fun correlation coefficient and WM-Fun correlation coefficient for every individual. The absolute value of correlation coefficient for each individual was used to measure the strength of network coupling.

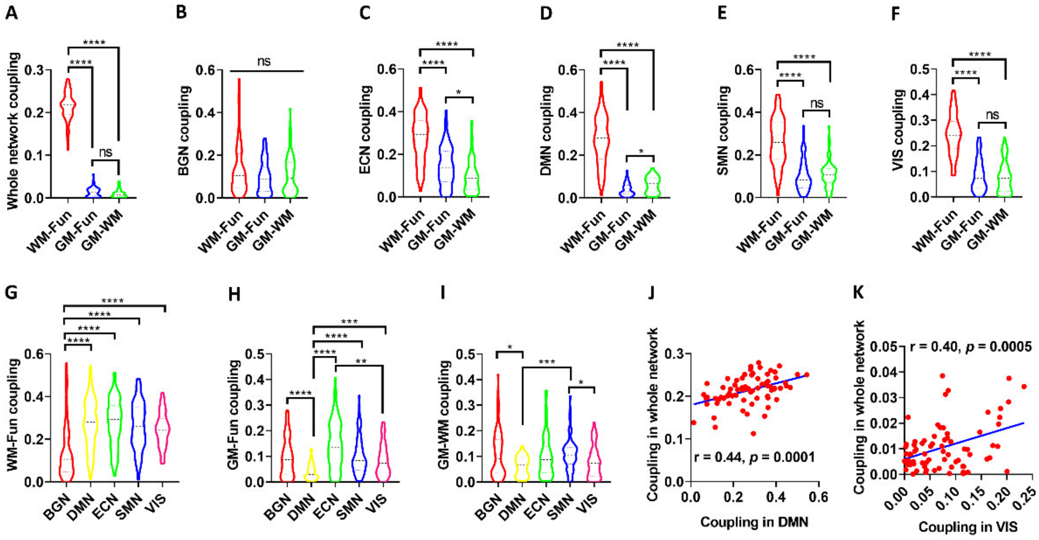


Supplementary Fig. 2. Network couplings varied among different network types. Strength comparisons of WM-Fun network coupling with GM-Fun network coupling and GM-WM network coupling in the whole network (A), BGN (B), DMN (C), ECN (D), SMN (E), and VIS (F). WM-Fun network coupling in BGN was much lower than that of other subnetworks, including DMN, ECN, SMN, and VIS (G). GM-Fun network coupling in DMN was lower than that of other networks, including BGN, ECN, SMN, and VIS (H). GM-WM network coupling in DMN was much lower than other subnetworks, including BGN and SMN (I). WM-Fun network coupling strength in DMN was significantly correlated with WM-Fun network coupling strength in the whole network (J). GM-WM network coupling strength in VIS was significantly correlated with GM-WM network coupling strength in the whole network (K). Nonparametric Kruskal-Wallis test following post hoc Dunn's test was used to compare the difference of network coupling strengths. Pearson's correlation was used to analyze the relationships of network couplings. *$p < 0.05$, **$p < 0.01$, ***$p < 0.001$, ****$p < 0.0001$.

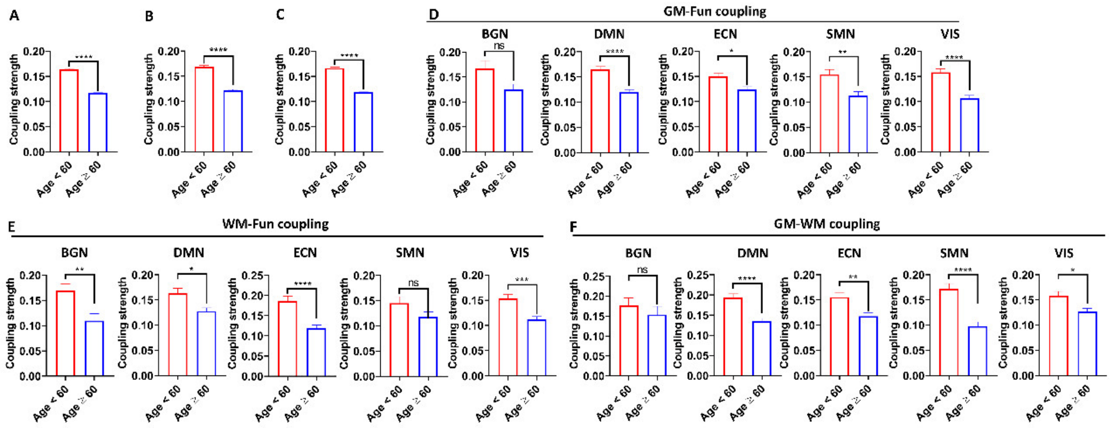


Supplementary Fig. 3. Reduced node couplings in PD patients with older age. Reduced strengths of GM-Fun node coupling (A), WM-Fun node coupling (B), and GM-WM node coupling (C) in the whole network of PD patients with age ≥ 60 (n = 48) compared to age < 60 (n = 25). Group difference of strengths of GM-Fun node coupling (D), WM-Fun node couplings (E), and GM-WM node coupling (F) in 5 subnetworks. Nonparametric Mann-Whitney U test was used to compare the difference of node coupling strengths between patients with age ≥ 60 and patients with age < 60. *$p < 0.05$, **$p < 0.01$, ***$p < 0.001$, ****$p < 0.0001$.

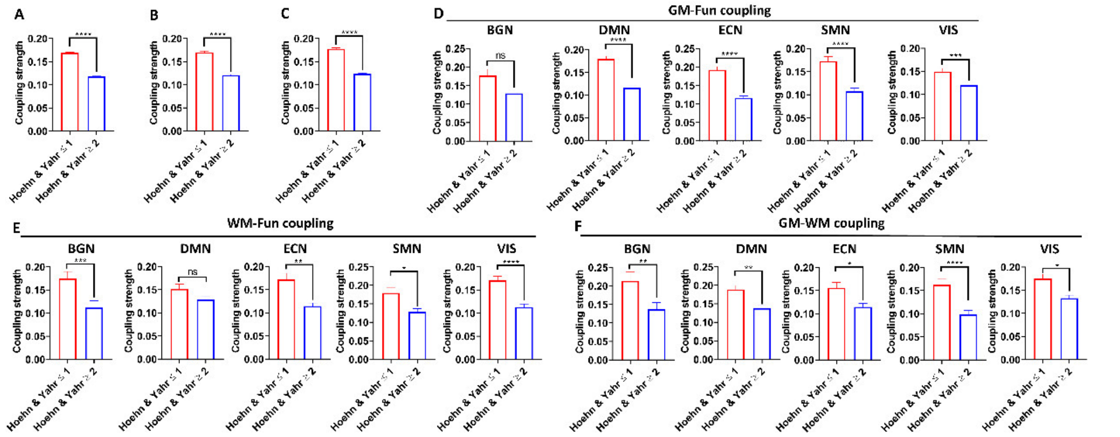


Supplementary Fig. 4. Reduced node couplings in PD patients with more severe disease. Reduced strengths of GM-Fun node coupling (A), WM-Fun node coupling (B), and GM-WM node coupling (C) in the whole network of PD patients with Hoehn & Yahr stage ≥ 2 (n = 50) compared to Hoehn & Yahr stage ≤ 1 (n = 23). Group difference of strengths of GM-Fun node coupling (D), WM-Fun node couplings (E), and GM-WM node coupling (F) in 5 subnetworks. Nonparametric Mann-Whitney U test was used to compare the difference of node coupling strengths between patients with Hoehn & Yahr stage ≥ 2 and patients with Hoehn & Yahr stage ≤ 1. $*p < 0.05$, $**p < 0.01$, $***p < 0.001$, $****p < 0.0001$.

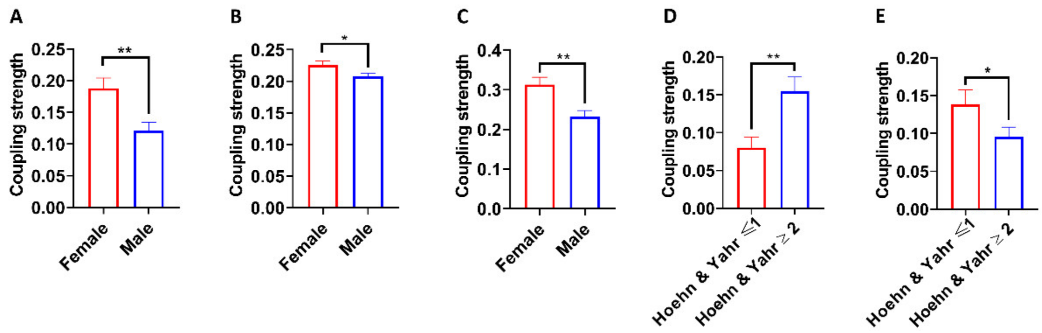


Supplementary Fig. 5. The effects of sex and disease severity on network coupling of PD patients. Male patients (n = 47) exhibited lower strength of GM-Fun network coupling in ECN (A) and lower strength of WM-Fun network coupling in the whole network (B) and SMN (C) compared to female patients (n = 26). Patients with Hoehn & Yahr stage ≥2 (n = 50) showed higher WM-Fun network coupling in BGN (D) and lower GM-WM network coupling in BGN (E) compared to patients with Hoehn & Yahr stage ≤1 (n = 23). Nonparametric Mann-Whitney U test was used to compare the difference of network coupling strengths. *$p < 0.05$, **$p < 0.01$.

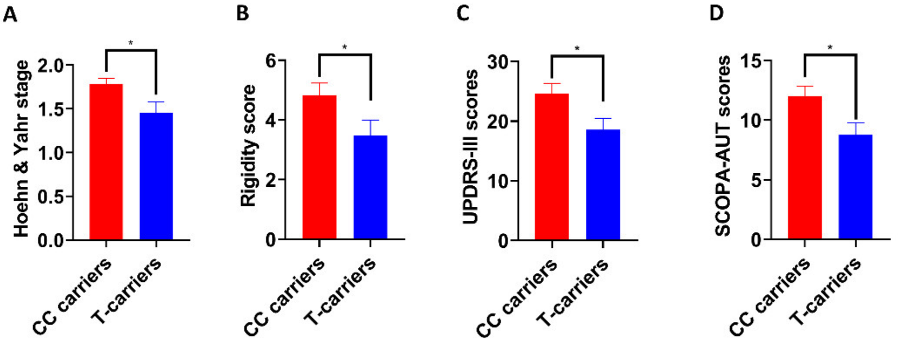


Supplementary Fig. 6. *TMEM175* rs34311866 CC carriers exhibited more severe motor and non-motor symptoms compared to T-carriers. Group difference (n = 48 for CC carriers, n = 25 for T-carriers) of Hoehn & Yahr stage (A), Total Rigidity scores (B), UPDRS-III scores (C), and SCOPA-AUT scores (D). Unpaired t-test was used to compare the difference of scores of clinical scales. *$p < 0.05$.
